# Case- fatality rate in COVID- 19 patients: A meta- analysis of publicly accessible database

**DOI:** 10.1101/2020.04.09.20059683

**Authors:** Souvik Maitra, Mansij Biswas, Sulagna Bhattacharjee

**Affiliations:** Department of Anaesthesiology, Pain Medicine & Critical Care, All India Institute of Medical Sciences, New Delhi; Medical Advisor, Boehringer Ingelheim India

**Keywords:** COVID-19, SARS-CoV-2, novel coronavirus, epidemiology, mortality rate

## Abstract

A novel coronavirus was reported in Wuhan, China in December 2019 to cause severe acute respiratory symptoms (COVID-19). In this meta-analysis, we estimated case fatality rate from COVID-19 infection by random effect meta-analysis model with country level data. Publicly accessible web database WorldOMeter (https://www.worldometers.info/coronavirus/) was accessed on 24th March 2020 GMT and reported total number of cases, total death, active cases and seriously ill/ critically ill patients were retrieved. Primary outcome of this meta-analysis was case fatality rate defined by total number of deaths divided by total number of diagnosed cases. Pooled case fatality rate (95% CI) was 1.78 (1.34-2.22) %. Between country heterogeneity was 0.018 (p<0.0001). Pooled estimate of composite poor outcome (95% CI) was 4.06 (3.24-4.88) % at that point of time after exclusion of countries reported small number of cases. Pooled mortality rate (95% CI) was 33.97 (27.44-40.49) % amongst closed cases (where patients have recovered or died) with. Meta regression analysis identified statistically significant association between health expenditure and case fatality rate (p=0.0017).

## Background

A novel coronavirus was reported in Wuhan, China in December 2019 to cause severe acute respiratory symptoms (COVID-19) [1]. Subsequently, this viral outbreak was reported in 197 countries and one international ship on 24^th^ March 2020. A wide range of mortality rate was reported from this viral illness and World Health Organization reported mortality rate of 3.4% on 3^rd^ March 2020 [2].

## Objectives

In this meta-analysis, we estimated ‘case fatality rate’ from COVID-19 infection by random effect meta-analysis model with country level data.

## Methods

Publicly accessible web database *‘WorldOMeter’* (https://www.worldometers.info/coronavirus/) was accessed on 24^th^ March 2020 GMT and reported total number of cases, total death, active cases and seriously ill/ critically ill patients were retrieved. Primary outcome of this meta-analysis was ‘case fatality rate’ defined by total number of deaths divided by total number of diagnosed cases. Secondary outcomes were mortality rate amongst closed cases (where patients recovered or died) and proportion of composite poor outcome (defined by the number of patients died or critically ill). Meta-regression analysis was performed to identify association between population density, health expenditure (percentage of GDP) and percentages of patients over 65y age with mortality. Between country heterogeneity (τ) was estimated by restricted maximum likelihood method and 95% prediction interval of pooled mortality were reported [3]. Countries reporting small number of cases (<100 on 24^th^ March 2020) were excluded from analysis; however, sensitivity analysis was planned with including them. Association between mortality and clinical characteristics of the patients were assessed by mixed effect meta-regression model.

All analyses were conducted in *metafor* package in R (R version 3.6.1; R Foundation for Statistical Computing, Vienna, Austria).

## Results

At the time of database access, 422582 patients were infected with COVID-19 with 18891 patients have died till that point of time. At that point of time 13269 patients were critically or seriously ill. Pooled case fatality rate (95% CI) was 1.78 (1.34-2.22) % with I^2^= 99.18%. Between country heterogeneity (τ) was 0.018 (p<0.0001). Sensitivity analysis was performed by exclusion of one country at times and pooled estimates of crude fatality rate remained mostly similar. Another sensitivity analysis was performed including countries where reported total number of cases were less than 100, and estimated case fatality rate (95% CI) was 1.77% (1.44-2.14) % with I^2^= 97.75%. Significant amount of publication bias was identified by regression test both with (p<0.0001) and without (p=0.0017) including countries will small number of cases. Pooled estimate of composite poor outcome (95% CI) was 4.06 (3.24-4.88) % with I^2^= 99.44% at that point of time after exclusion of countries reported small number of cases.

Pooled mortality rate (95% CI) was 33.97 (27.44-40.49) % amongst closed cases (where patients have recovered or died) with I^2^=99.88% [figure 1]. Meta regression analysis identified statistically significant association between health expenditure and case fatality rate (p=0.0017) [figure 2].

**Figure 1:**
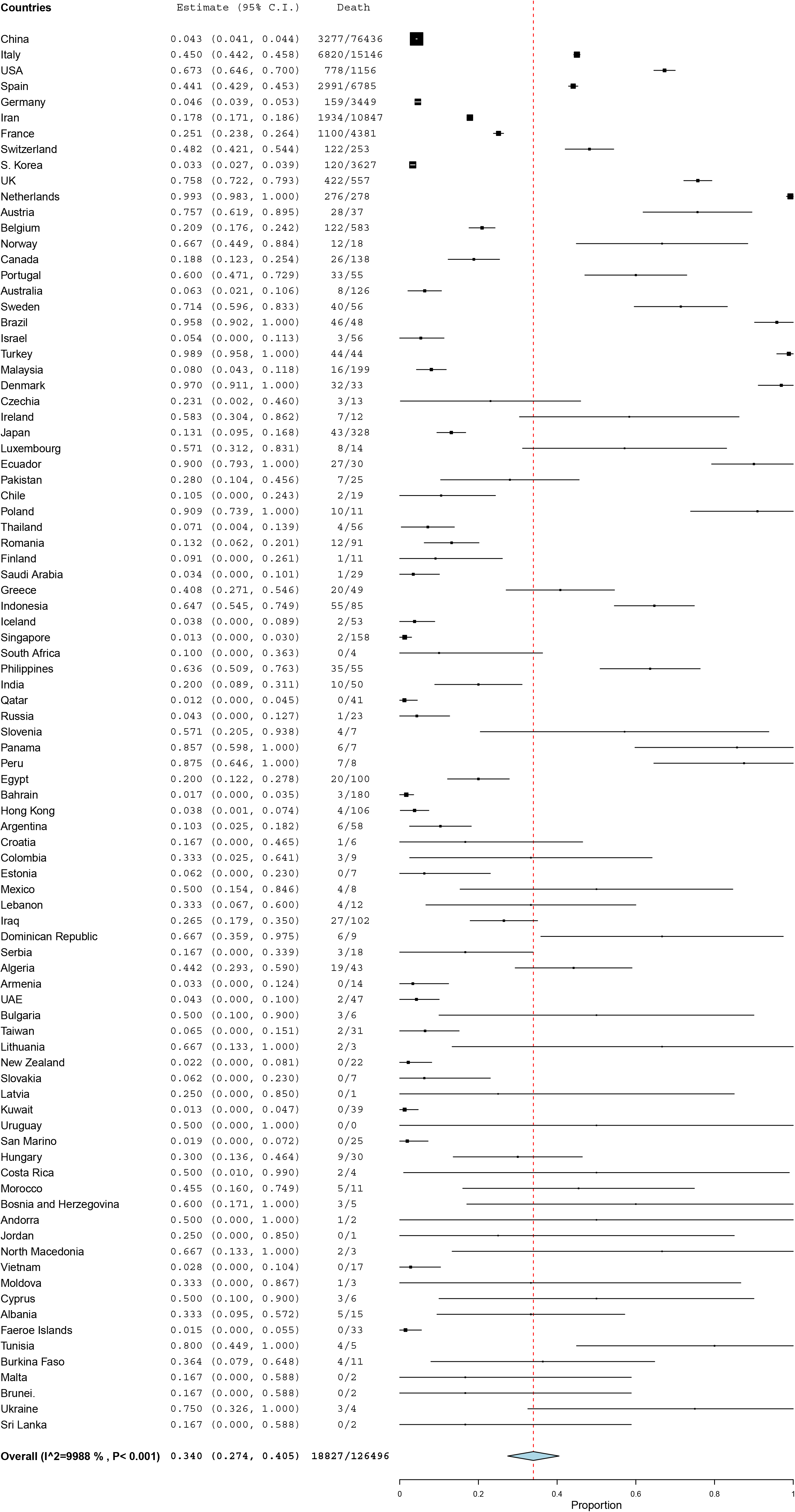
Forest plot showing estimated mortality rate amongst closed cases at country level and pooled estimate of mortality rate.

**Figure 2:**
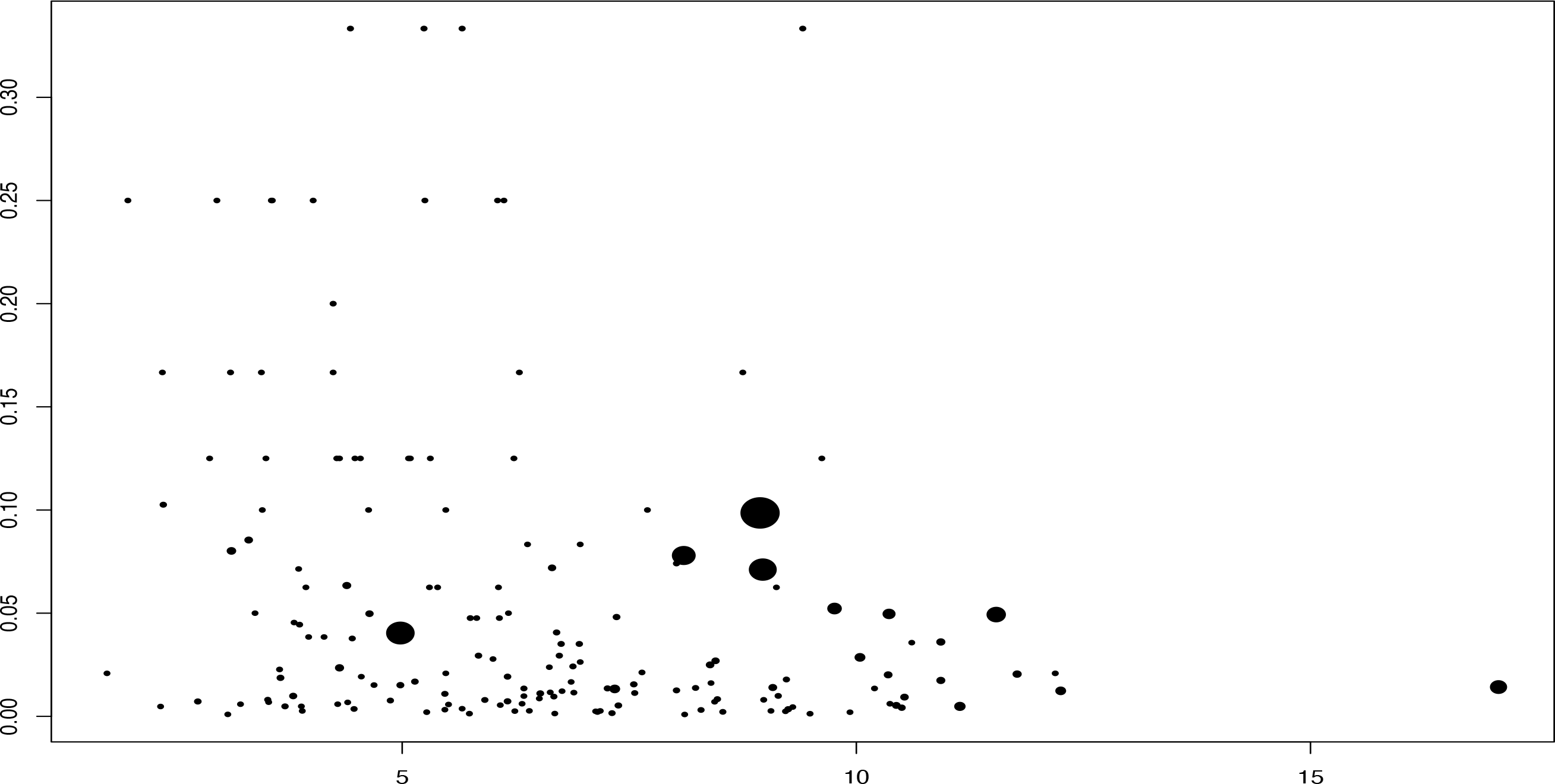
Bubble plot showing association between health expenditure (expressed as percentages of GDP) and log of case fatality rate.

## Discussion

We have found a pooled mortality rate of 1.77%, which was less than WHO reported death rate of 3.4% but similar to the first published report of mortality from Wuhan, China. [1] Our pooled estimate of composite poor outcome was around 4%, which was similar to the previous report from China. Another important finding is that presence of a significant heterogeneity and publication biases in the pooled analysis. Underreporting from different countries and heterogenous testing strategy and quality of care are probably reason thereof. Estimation of mortality rate at the time of outbreak is difficult because there are always a proportion of newly diagnosed cases where clinical outcome (death or recovery) is yet to happen. Secondly, total number of diagnosed cases depends upon the diagnostic strategy of the country and proportion of asymptomatic cases. Surprising high mortality were obtained when only closed cases were used in denominator; however, true mortality rate is expected to be smaller as a large number of suspected patients were not subjected to laboratory confirmation. So, actual mortality from COVID-19 is difficult to estimate at this point of time and it could higher than current estimation.

## Data Availability

Data will be available from the authors

## References

1. Guan WJ, Ni ZY, Hu Y, et al. Clinical Characteristics of Coronavirus Disease 2019 in China [published online ahead of print, 2020 Feb 28]. N Engl J Med.

2. WHO Director-General’s opening remarks at the media briefing on COVID-19 - 3 March 2020 - World Health Organization, March 3, 2020

3. Deeks JJ, Higgins JPT, Altman DG (editors). Chapter 9: Analysing data and undertaking meta-analyses. In: Higgins JPT, Green S (editors). Cochrane Handbook for Systematic Reviews of Interventions Version 5.1.0 (updated March 2011). The Cochrane Collaboration, 2011. Available from www.handbook.cochrane.org.

